# Increased burden of endogenous viral elements in Alzheimer’s Disease compared to cognitively healthy centenarians

**DOI:** 10.64898/2026.10.01.26364419

**Authors:** E. Ma, N. Tesi, A. Salazar, M. Hulsman, H. Holstege, M.J.T. Reinders

## Abstract

**INTRODUCTION:** Throughout history, viruses have been infecting humans, leaving genomic traces called endogenous viral elements (EVEs). This study explores the infectious hypothesis for Alzheimer’s disease (AD) that suggests that viral infections may contribute to AD by analyzing EVEs in genomes of AD patients and cognitively healthy centenarians (CHCs) measured by long-read sequencing.

**METHODS:** We implemented an EVE detection pipeline that screened the genomes of 248 AD patients and 240 CHCs. EVEs were measured based on EVE count, length, and chromosome location.

**RESULTS:** The genomes of AD patients contain significantly more EVEs than those of CHCs (p=3.2×10^-4^). Moreover, certain chromosomal regions in AD patients show a higher EVE count, particularly of the Orthoherpesviridae family, near genes differentially expressed in AD brains.

**DISCUSSION:** These findings suggest that viral infections may increase susceptibility to AD, highlighting that preventive measures against infections potentially reduce AD risk.

**Research in context:** *Systematic review:* Alzheimer’s disease (AD) is classically linked to β-amyloid accumulation, but β-amyloid may also act as an antimicrobial peptide against viruses like herpesvirus, supporting an infectious hypothesis where viral infection triggers β-amyloid production. We tested this using endogenous viral elements via long-read sequencing in AD patients and cognitive healthy centenarians (CHC).

*Interpretation:* AD genomes carry significantly more endogenous viral elements than CHC genomes, with families including *Orthoherpesviridae* enriched at chromosomal regions near genes differentially expressed in AD brains and linked to a herpesvirus infection pathway.

*Future directions:* Our computational analyses have identified novel human genetic loci that may influence Alzheimer’s disease pathology, opening new avenues for in vivo and in vitro investigations into its underlying mechanisms. Additionally, we emphasize the significance of endogenous viral elements in Alzheimer’s research and advocate for a global exploration of viral element heterogeneity to deepen our understanding of their potential role in disease progression.

**Highlights:**

- First long-read screen for EVEs comparing AD patients and centenarians.
- AD genomes carry significantly more endogenous viral elements than CHCs.
- Chr19 herpesvirus-enriched genes map to the Herpes Simplex Virus 1 pathway.

## 1. Background

Alzheimer’s disease (AD) is the most common form of dementia and imposes a major socio-economic burden on society and patients’ families [1]. The most widely accepted theory describing the molecular events leading to AD is the amyloid hypothesis, which proposes that AD is primarily caused by the accumulation of β-amyloid (Aβ) plaques in the brain [2]. These plaques are thought to trigger neuroinflammation, oxidative stress, and synaptic dysfunction, ultimately leading to neuronal loss [2].

However, recent evidence suggests that Aβ may also function as an antimicrobial peptide (AMP) rather than being only a mere byproduct of protein metabolism [3]. Specifically, Aβ shows striking structural and functional similarity to the antimicrobial LL-37 peptide [3] and has been conserved for over 400 million years across the animal kingdom [4]. Aβ inhibits the growth of various pathogens, including influenza A virus *in vitro* [5] and Herpes Simplex Virus-1 *in vivo* [6], and was shown to co-localize with herpesvirus particles in the brains of AD patients more frequently than in healthy controls [7]. These findings support the infectious hypothesis, which suggests that viral infections contribute to AD pathogenesis by triggering Aβ production in response to real or perceived infections.

Throughout evolution, humans have been exposed to a wide range of viral infections, some of which have left genomic traces, known as endogenous viral elements (EVEs) [8]. These viral remnants, integrated into the genome, constitute 5-8% of the human genome, with the majority classified as human endogenous retroviruses (HERVs), while others belong to non-retroviral EVEs [9]. Viruses integrate into the host genome through different mechanisms: retroviruses such as HIV and HERVs use a specialized enzyme called reverse transcriptase to convert their RNA genome into DNA. This DNA is then inserted into the host genome by another viral enzyme, integrase, creating a provirus that can be stably inherited through generations if it affects germline cells. In contrast, non-retroviral viruses (such as some DNA viruses) can integrate into the genome by random recombination events, often through double-strand break repair mechanisms. While some EVEs have become genetically inert, others have been co-opted by the host for essential functions, such as the *Syncytin* gene in placental development [10, 11]. However, not all integrated viral sequences are beneficial. In fact, some may retain pathogenic potential, and may be transcriptionally reactivated under certain conditions such as inflammation [12], autoimmune diseases [13], or cancer [14].

From a genetic perspective, EVEs can vary significantly in size depending on the type of virus and the extent of degradation over evolutionary time. While some elements are fragmented, many span several kilobase pairs (kbp), necessitating long-read sequencing and advanced assembly methods to resolve their structural complexity.

Here, we systematically investigated the infectious hypothesis of AD from an endogenous perspective. We screened long-read sequencing-based genome assemblies from AD patients and Cognitively Healthy Centenarians (CHCs) for differences in viral content, using the non-redundant Refseq viral protein database [15].

## 2. Methods

### 2.1 Population of the study

We included 248 individuals diagnosed with AD from the Amsterdam Dementia Cohort (ADC) [16]. We contrasted these to 240 CHCs from the 100-plus Study [17]. A detailed description of all individuals included is available elsewhere [18, 19]. All participants and/or their legal representatives provided informed consent for participation in clinical and genetic studies. Blood samples of all AD and CHC individuals were processed and assessed for minimum quality requirements as previously described [18–20]. All samples were long-read sequenced using PacBio Sequel IIe instrument with at least 1 SMRT cell. Whole-genome phased de novo assembly was performed with hifiasm (with --n-hap 2 parameter). Assembled contigs were then aligned to GRCh38 using minimap2 (with-ax asm10 parameter) [21, 22]. The complete data processing pipeline is freely available at https://github.com/holstegelab/snakemake_pipeline. We then used aligned and un-aligned assembled contigs relative to the paternal and maternal haplotypes for downstream analyses.

### 2.2 Overview of the detection pipeline

An overview of the detection pipeline is shown in Figure 1. Briefly, we: (1) screened assembled contigs for subsequences resembling viral proteins using a local alignment algorithm; (2) merged overlapping viral-like alignments within the same region, excluding those showing significant similarity to human proteins; (3) identified the genomic coordinates of the resulting viral integrations; (4) discarded regions that were situated within non-viral structures of the human genome and annotated the remaining regions; and (5) performed downstream analyses to investigate differences in viral content between the AD and CHC cohorts. The detection pipeline is freely available at https://github.com/holstegelab/Eduard_Ma_Master_thesis.

**Figure 1:**
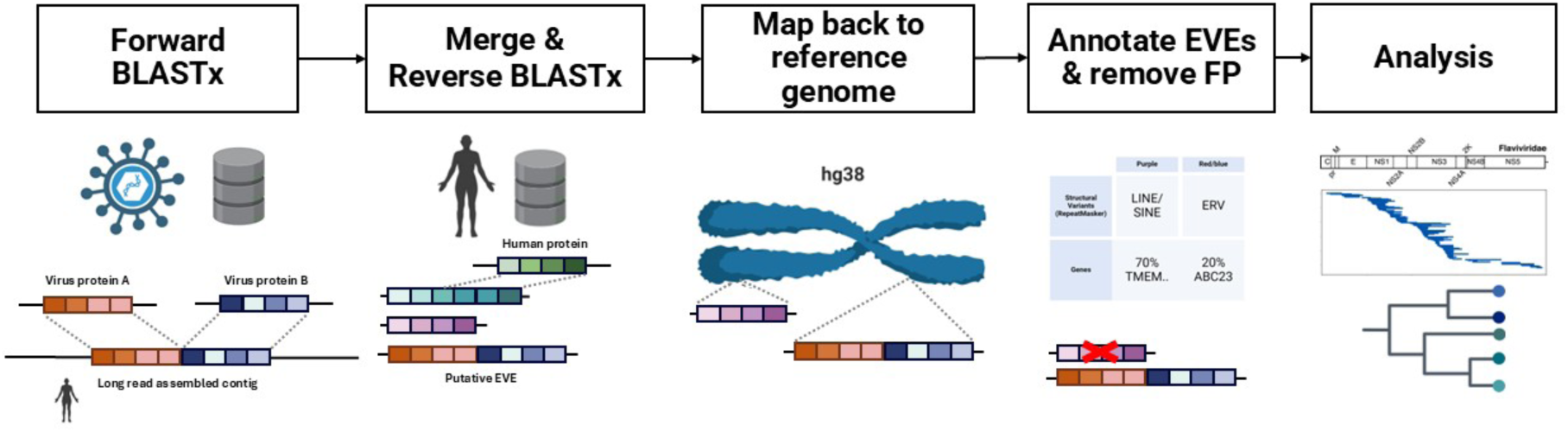
The EVE detection pipeline. From left to right: 1) Human sequences were queried against the Refseq virus database; 2) The putative EVEs were queried against a human protein database and the suspect EVEs that resembled human sequences were discarded; 3) The contig coordinates were mapped back to positions on human chromosomes; 4) Further filtering was performed to exclude putative EVEs that overlapped known repetitive regions in the genome, such as LINEs and SINEs; 5) Annotation of the candidate EVE regions.

### 2.3 EVE detection – forward screening

We first identified candidate EVEs by performing a BLASTx search, specifying our AD and CHC phased assemblies (in FASTA format) as the query sequences, and the non-redundant RefSeq NCBI library database as the database [23] (https://ftp.ncbi.nih.gov/refseq/release/viral/, accessed March 2024, 683,238 protein sequences) [24–26]. We refer to this as the *forward* BLASTx search. DIAMOND BLASTx was used as it is significantly faster than NCBI’s default BLASTx algorithm [23]. The nucleotide query sequences (i.e. the human assembled genomes) were translated in six reading frames (resulting in six protein sequences) and were compared against the viral protein sequence database using default settings. Human subsequences with high similarity to viral protein sequences (e-value < 10^-6^ were extracted. Overlapping regions within the same genome were merged and the alignment with the lowest e-value was chosen as representative for that merged region. Nucleotide sequences of the putative endogenous viral elements (EVEs) were then extracted using BedTools [27]. This analysis was performed for each of the 488 individuals, and each of the maternal and paternal haplotypes.

### 2.4 EVE detection – reverse search

We further refined the list of candidate EVEs by removing those that were highly similar to human proteins. To do so, we performed another DIAMOND BLASTx alignment, this time using the human protein database as input (https://www.ncbi.nlm.nih.gov/genome/guide/human/, accessed January 2024) [23]. Once again, the candidate EVEs regions were translated into protein sequences, which were then compared to all human protein sequences. We refer to this as the *reverse* BLASTx search. Any subsequence that was highly similar to a human protein (e-value < 10^-6^) and overlapped for at least 50% with that human protein was considered a false positive and excluded from any downstream analysis. Of note, alignments to human proteins known as “endogenous viral” were excluded in our reverse BLASTx filtering step. We then identified the chromosomal locations of candidate EVEs in the space of the GRCh38 reference genome. To do so, for each candidate EVE, we derived the alignment positions to GRCh38 by parsing the CIGAR string of the relative contigs.

### 2.5 False positive removal and annotation

The candidate EVEs may still have significant homology with repetitive regions in the human genome that cannot be attributed to viral elements. Therefore, we used RepeatMasker annotations relative to GRCh38 to identify repeat annotations that overlapped the set of candidate EVEs [28]. Candidate EVEs that overlapped for more than 50% with an annotated human sequence were considered false positive, except for sequences that RepeatMasker itself classified as “ERV” or “Borna-like”. Furthermore, we retained only virus families known to integrate in vertebrate species as reported by the International Committee on Taxonomy of Viruses (ICTV) Master species list (ICTV_Master_Species_List_2023_MSL39.v1.xlsx, https://ictv.global/msl, accessed May 2024). Of note, *Metaviridae*, classified as a vertebrate virus family in ICTV, but being truly a collection of retrotransposons (i.e. Ty3/Gypsy Long-Terminal Repeat (LTR) retroelements), was included in the analyses [29]. Lastly, we annotated the refined set of EVEs using the NCBI taxonomy database and annotated EVEs to be within a gene or being intergenic based on their intersection with gene coordinates using BedTools intersection with default parameters (https://ftp.ncbi.nih.gov/pub/taxonomy/accession2taxid/, accessed March 2024) [27].

### 2.6 Statistical analyses

We tested whether the number of EVEs was significantly different between AD individuals and CHCs using negative binomial regression analysis (NBR) [30]. Similarly, the average lengths of EVEs were compared between AD individuals and CHCs using the two-tailed Mann-Whitney U test (MWU) [31]. Differences in proportions between the two cohorts were tested using the two-tailed Fisher’s exact test (FE) [32]. Where multiple tests were involved, we corrected p-values following the Benjamini-Hochberg procedure (BH), with a significance threshold set at 0.05 [33].

### 2.7 Implementation

The EVE detection pipeline was implemented as a mixture of bash and python scripts. All figures were created using matplotlib, Seaborn, Pygenometracks, and ETE Toolkit plotting packages [34–37]. Statistical analyses were performed through Scipy and PyMC statistical software packages [38, 39]. Wherever gene set enrichment analysis was performed, gProfiler was used with standard settings and based on Ensembl 113 and Ensembl Genomes 59 [40].

## 3. Results

### 3.1 Demographics and sequencing

The genomes of 248 individuals diagnosed with AD and 240 CHCs were sequenced using PacBio long-read technology and preprocessed as described previously (Methods). The majority of the participants in the study were female (Figure 2A). Overall, the AD genomes were sequenced at a median coverage of 18.2x, with median read-lengths of 14.8 kbp, and the CHC genomes were sequenced at a median coverage of 20.1x, with median read lengths of 14.7 kbp (Figure 2B).

**Figure 2:**
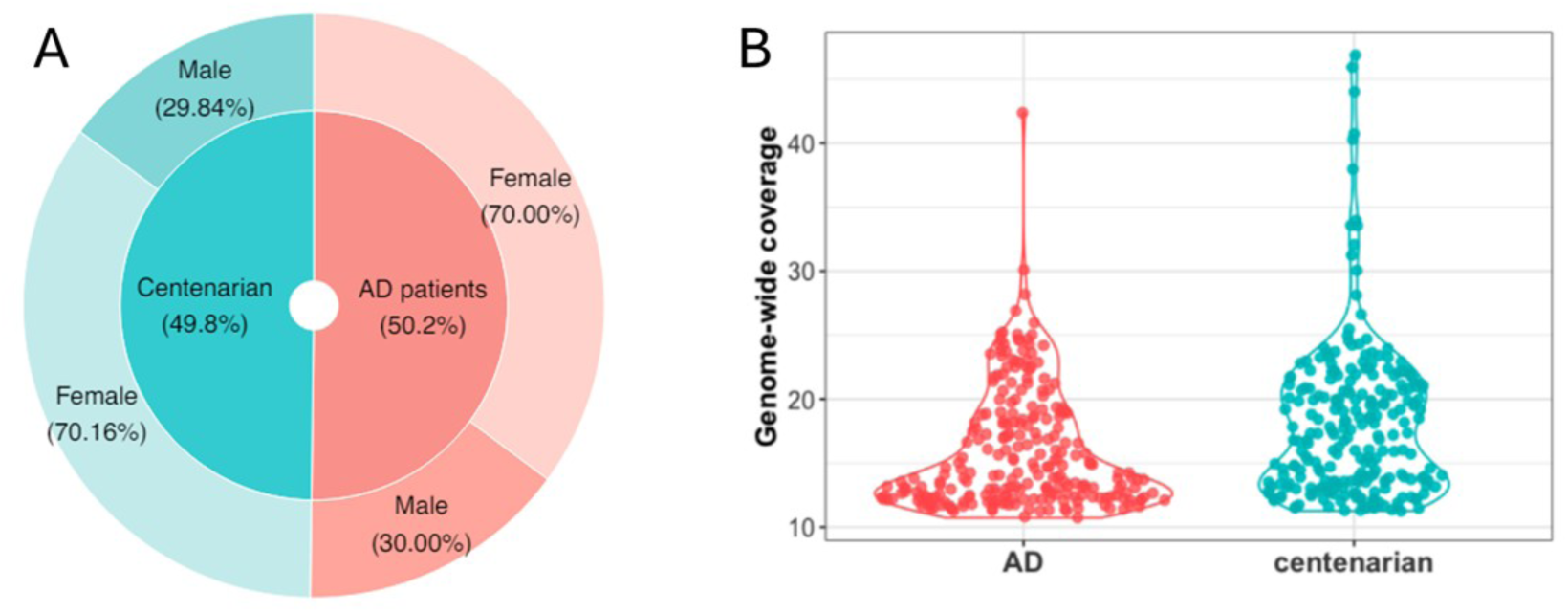
100+ study data characteristics outlining the relative sizes of the two cohorts and their sex proportions (A) and the distribution of genome coverage for the samples of both cohorts (B).

### 3.2 AD genomes harbor more EVEs

Numerous EVEs were identified in the genomes of both CHCs and AD patients. On average, 3,740 (± 114) EVEs were found in AD genomes, whereas 3,166 (±118) were found in CHCs. Consequently, the genomes of AD patients have a higher number of EVEs than the CHCs (p = 3.2×10^-4^, NBR). In total, subsequences similar to eleven viral families that are known to infect vertebrates were identified, including (+)RNA viruses *Retro*-, *Flavi*-, and *Metaviridae*, (-)RNA viruses *Bornaviridae*, ambisense RNA virus *Arenaviridae*, as well as dsDNA viruses *Orthoherpes*-, *Alloherpes*-, *Pox*-, *Irido*-, *Adeno*- and *Adintoviridae*.

All individuals in both cohorts carried viral integrations of the Retroviridae family, as expected given the obligatory integration step during infection (Figure 3A). EVEs of the *Borna*-, *Orthoherpes* and *Poxviridae* families were discovered in more than 90% of the individuals of both cohorts. Strikingly, we observed a significantly higher number of AD individuals with a *Flaviviridae* EVE in their genome (p = 3.8×10^-3^, FE). The remaining viral families were found sporadically in less than 50% of the individuals, for each cohort.

**Figure 3:**
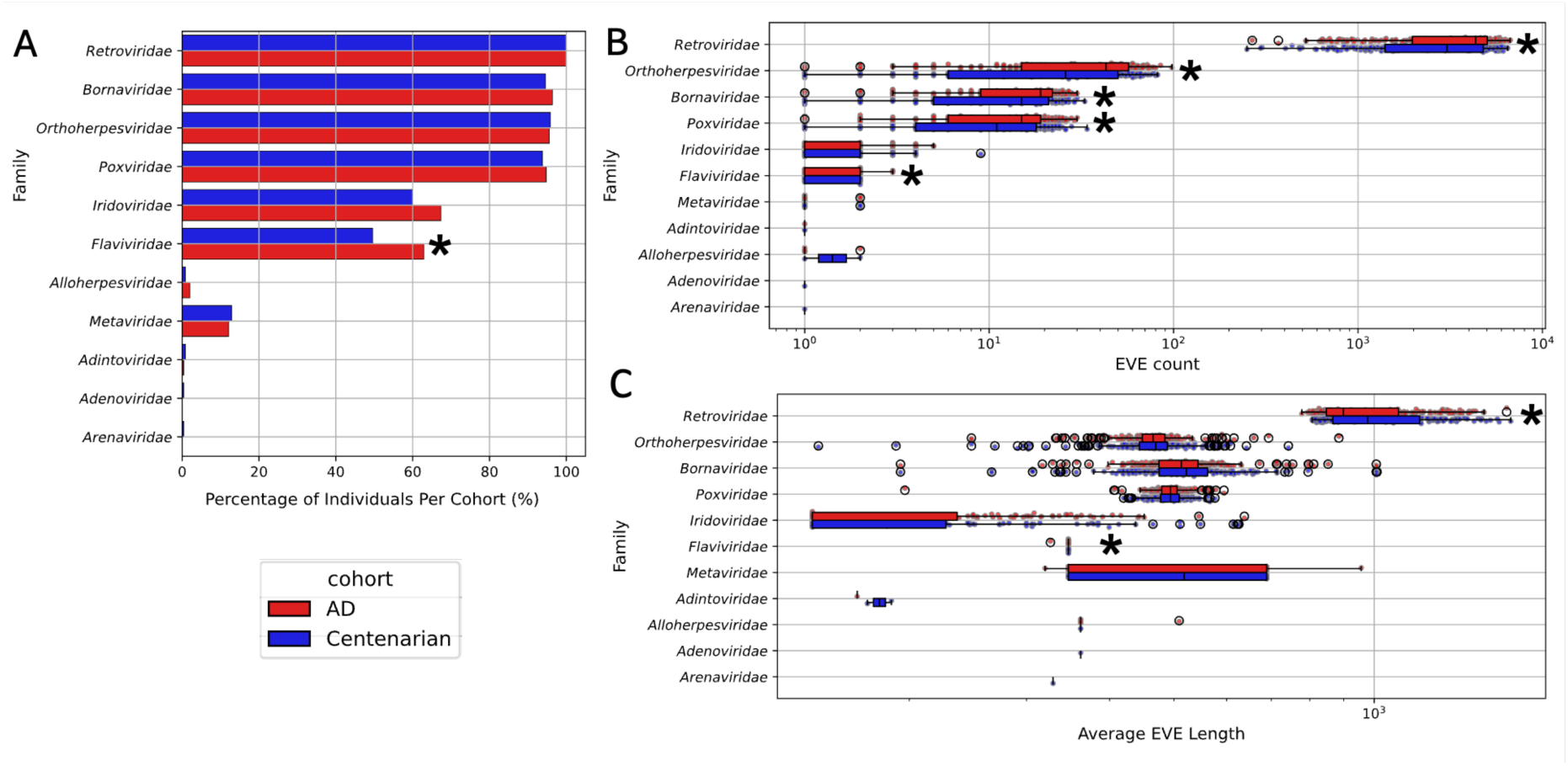
Percentage of individuals that have at least one EVE for the indicated virus family in their genome discovered, for the AD cohort (red) and cognitive healthy centenarian cohort (blue). B) Number of EVEs discovered for the indicated virus family per individual per cohort. Every dot represents an individual. C) EVE length distribution per individual per virus family with every dot representing one individual. Star markers indicate statistically significant differences between the cohorts.

We observed a general pattern of AD genomes that harbor significantly more EVEs of the *Retro*-, *Orthoherpes*-, *Borna*-, *Pox*-, and *Flaviviridae* families (FDR = 1.3×10^-3^, 1.3×10^-3^, 1.3×10^-3^, 3.1×10^-3^, 1.3×10^-3^ respectively, NBR, Poisson) (Figure 3B). The number of *Retroviridae* EVEs was in the thousands, whereas the other non-retroviral families were present in the order of tens, which is in line with the current knowledge of EVEs [8].

Lastly, we compared the average EVE length (bp) of the viral families per individual between AD and CHC. We found that the average EVE length of AD genomes was smaller than in CHCs for Retro-and Flaviviridae (FDR=0.01 and 0.02, respectively) (Figure 3C). This suggests that while AD genomes generally carry a higher number of EVE insertions, the average insertion length is smaller than in CHCs.

### 3.3 Viral families are biased towards specific chromosomal regions in our genome

To get a more localized view on where these viral regions were situated, we analyzed the distribution of viral regions across the genome (Figure 4A-B). We tested the frequency difference between the two cohorts per chromosome and per virus family. After multiple testing correction, four chromosomes were observed to have a significantly higher number of *Orthoherpesviridae* integrations in the AD cohort than in the CHC cohort (Figure 4C). Specifically, we observed a significantly larger fraction of AD individuals having a herpes integration on chromosome 11, 15, 17, and 19 (corrected p-value = 1.9×10^-2^, 5.1×10^-3^, 1.9×10^-3^, 4.9×10^-3^, respectively, FE). Additionally, all individuals have retroviral integrations on all chromosomes due to their abundant presence (the Y-chromosome was excluded from analyses). Remaining chromosomes seem to have similar integration frequencies between the two cohorts.

**Figure 4:**
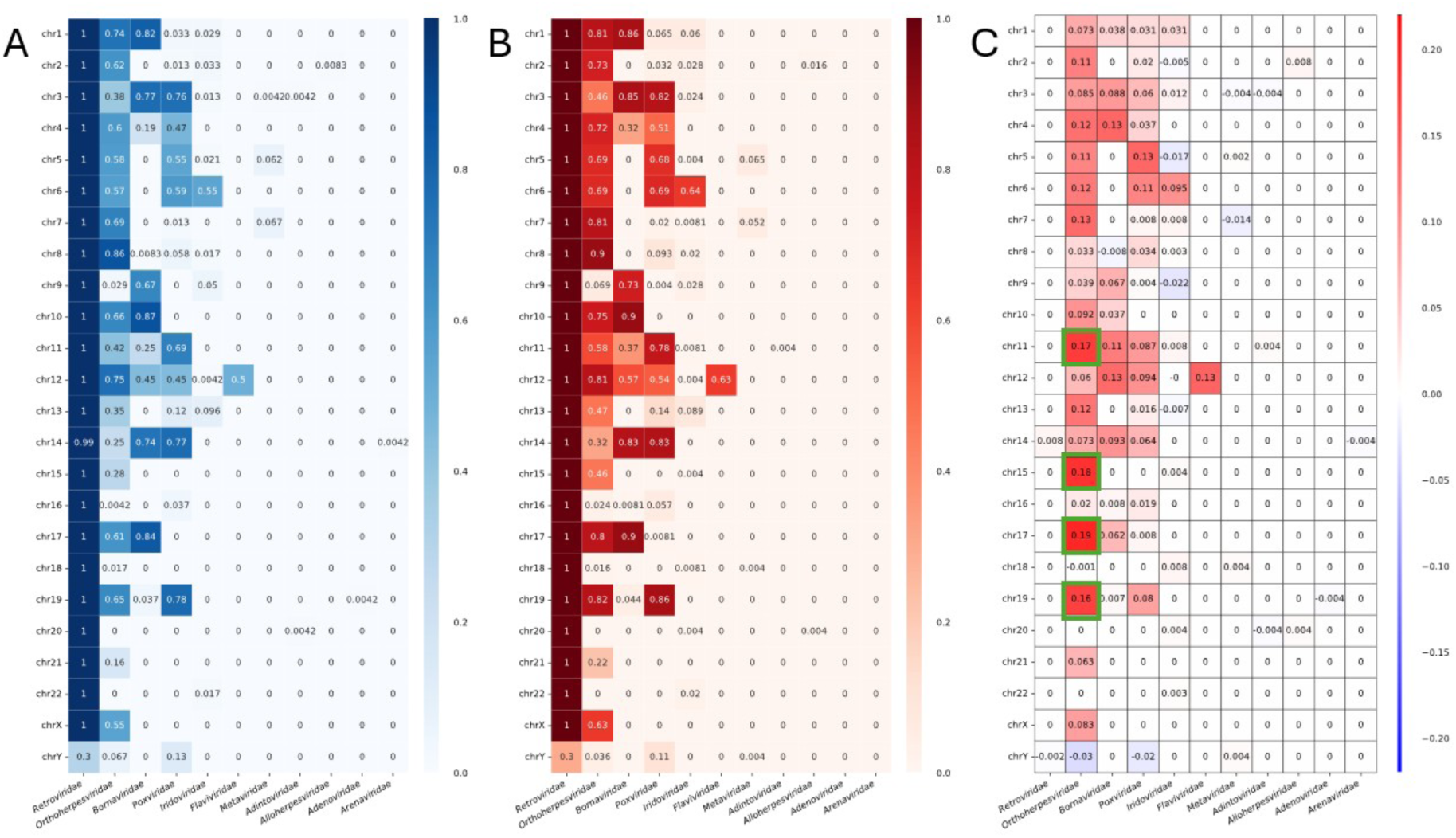
Per indicated viral family (columns), the number of individuals divided by the total cohort count that have at least one EVE per chromosome for (A) the CHC cohort, (B) the AD cohort. (C) The difference between fractions of (A) and (B). Highlighted in green are the chromosomes for which the herpes virus family showed statistically significant enrichments.

### 3.4 Viral integrations are localized to genes involved in herpes simplex infection

To better understand the chromosomal differences, we identified overlapping viral segments across all samples and counted how many individuals had an EVE in those segments (Methods). For five regions, we observed significantly more individuals in the AD group having an EVE compared to the CHC group, with the AD group always having higher counts (Table 1 and Figure 5C). One of such regions on chromosome 11 was located inside an LTR region of HERV3-int and overlapped with *RP11-75G1.6* (ENSG00000250508), a long-non coding RNA that is over-expressed in Parkinson’s disease [41]. We could not identify a significant relationship to AD of the nearby genes.

**Figure 5:**
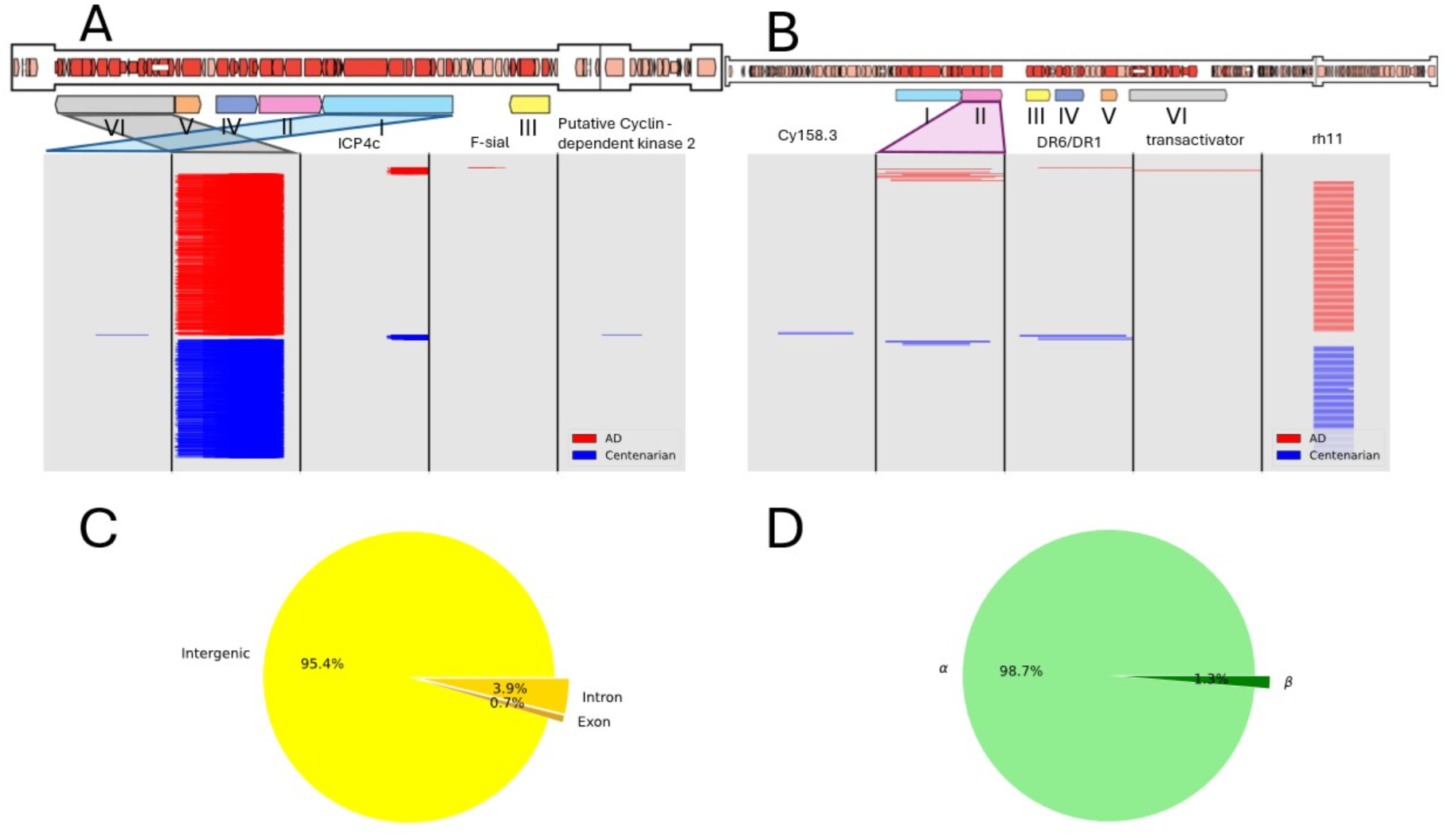
Coverage plot of Herpesviridae sequences in representative figures of the α-herpesvirus and β-herpesvirus genome in A and B, respectively. Roman letters indicate the six conserved domains in the herpesvirus genome: (I) Uracil-DNA glycosylase (II) Helicase-primase (III) DNA packaging terminase (IV) Major capsid protein (V) DNA polymerase catalytic subunit (VI) Envelope glycoproteins. Proteins that map back to non-conserved regions are annotated in the figure. Rough estimates based on literature show where they come from. Cy158.3 could not be mapped back. Schematics of the genomes were taken and adapted from [46]. C) Proportions of intergenic, exonic, and intronic herpes EVEs in the two cohorts D) Proportions of α- and β-herpesvirus found in the two cohorts.

**Table 1:** Overview of the significantly enriched regions for the *Orthoherpesviridae* virus family. Every region is specified through a chromosome number (Chr) and start and stop coordinates (Coordinates). *N_AD_* represents the number of AD individuals in which the specified enrichment has been found. Similarly *N_CHC_* represents the number of CHCs for which the enrichment was found. P-values (p) and odds ratio (OR) are reported for the FE tests after BH correction. UCSC genome browser overview for every chromosome showcases overlapping regions with other structural elements like LTRs (Region) [48, 49].

| Chr | Coordinates | $N_{AD}$ | $N_{CHC}$ | OR | p | Region |
| --- | --- | --- | --- | --- | --- | --- |
| 11 | 68873390-68873754 | 124 | 80 | 2 | $2.3 \times 10^{-4}$ | Suppl. Fig. 7 |
| 15 | 92751346-92751935 | 109 | 66 | 2.07 | $1.6 \times 10^{-4}$ | Suppl. Fig. 8 |
| 17 | 41067769-41068257 | 134 | 94 | 1.83 | $1.1 \times 10^{-3}$ | Suppl. Fig. 9 |
| 17 | 50413398-50413999 | 187 | 136 | 2.34 | $1.5 \times 10^{-5}$ | Suppl. Fig. 10 |
| 19 | 50347788-50348322 | 177 | 130 | 2.11 | $1.2 \times 10^{-4}$ | Suppl. Fig. 11 |

Similarly, regions on chromosome 15 and 17 that were dominant in herpes integrations showed overlap with LTR regions. On chromosome 15, one region overlapped with the *FAM174B* gene (ENSG00000185442), which has been shown to be downregulated in AD patients [42]. This downregulation may reflect local epigenetic repression associated with host-mediated silencing of the EVE, rather than reduced susceptibility to viral activity.

Finally, in chromosome 19, we observed a region to be overlapping with a LTR of the ERV1 family and *NR1H2* gene. Alterations in the expression of *NR1H2* have been proposed as a risk factor for AD [43]. Interestingly, we identified a set of genes on chromosome 19 having more herpes integrations in AD than in CHCs (minimum odds ratio of 1.5): *ZNF93, ZNF682, ZNF826P, ZNF737, ZNF626, ZNF66, ZNF708, ZNF98, ZNF91, RPSAP58, NAPSB, ZNF765, ZNF813*. More than half of these genes (*ZNF93, ZNF682, ZNF737, ZNF626, ZNF66, ZNF708, ZNF98, ZNF91*, *NAPSB, ZNF765, ZNF813)* were shown to be differentially expressed on the RNA level in at least one region of the brain based on the AMP-AD study (Table 2) [44]. Moreover, gene set enrichment analysis of these genes shows strong significance for multiple pathways, with Herpes simplex virus 1 infection pathways (KEGG:05168) being the most significant (p=3.2×10^-11^).

**Table 2:** Overview of genes that are differentially expressed on the RNA level in at least one region of the brain based on the AMP-AD study [45].

|  |  |
| --- | --- |
| ZNF93 | https://agora.adknowledgeportal.org/genes/ENSG00000184635 |
| ZNF682 | https://agora.adknowledgeportal.org/genes/ENSG00000197124 |
| ZNF737 | https://agora.adknowledgeportal.org/genes/ENSG00000237440 |
| ZNF626 | https://agora.adknowledgeportal.org/genes/ENSG00000188171 |
| ZNF66 | https://agora.adknowledgeportal.org/genes/ENSG00000160229 |
| ZNF708 | https://agora.adknowledgeportal.org/genes/ENSG00000182141 |
| ZNF98 | https://agora.adknowledgeportal.org/genes/ENSG00000197360 |
| ZNF91 | https://agora.adknowledgeportal.org/genes/ENSG00000167232 |
| NAPSB | https://agora.adknowledgeportal.org/genes/ENSG00000131401 |
| ZNF765 | https://agora.adknowledgeportal.org/genes/ENSG00000196417 |
| ZNF813 | https://agora.adknowledgeportal.org/genes/ENSG00000198346 |

### 3.5 The origin of herpes virus sequence

To better understand which viral proteins align well with human subsequences, we mapped the detected viral protein segments to a schematic version of the herpes genome. The majority of these segments were similar to the α-herpes envelope protein (Figure 5A-D. Other viral proteins were only sporadically found, with integrations mostly in intergenic regions (Figure 5D).

## Discussion

Ever since the first discovery of virus DNA inside the brains of AD patients, the so-called infectious hypothesis of AD has been under heavy dispute. True or not, the fact remains that virus infections have influenced and shaped our genome throughout human history. Evidence of these influences can be traced back by investigating our DNA for subsequences similar to virus proteins. Assuming that viruses might be an environmental contributory factor for the onset of AD, we hypothesized that traces of these virus integrations are to be found back more frequently in the genomes of AD patients. To test this hypothesis, we developed a BLASTx alignment-based pipeline to screen human assemblies constructed from PacBio long-read sequencing data against all known vertebrate viruses in the Refseq NCBI virus protein database [15]. With that, we provide a comprehensive perspective on the infectious hypothesis of AD by screening the high-quality genomes of AD patients and cognitive healthy centenarians (CHCs) for remnants of virus proteins from past infections.

We observed that AD genomes contained more EVEs than CHC genomes. We found that a higher fraction of AD patients carry an EVE of one of the three non-retroviral families, *Orthoherpes*-, *Borna*-, and *Poxviridae* on specific chromosomal locations compared to CHCs. Interestingly, while the number of EVEs in AD genomes was larger than in CHCs, the average EVE length was smaller. As with structural variants, however, biological impact may depend less on overall burden or size than on genomic context, including where EVEs occur and which genes or regulatory elements they may affect.

Specifically for the herpesvirus family, we found that EVEs tend to be close to genes that are differentially expressed in the brains of AD patients, compared to healthy control groups. We also observed that AD samples had relatively more herpesvirus EVEs on chromosome 19, and that the set of genes close to the herpesvirus EVEs on chromosome 19 is enriched for the “Herpes Simplex Virus 1 infection” pathway (KEGG:05168). When looking closer, the set of genes of the KEGG database play a role in the activity of Zinc Finger Antiviral Protein according to the KEGG database [45], with *ZAP* being an important protein for the cell’s intrinsic immune response [46].

Taken together, our data might suggest that individuals with more endogenous viral elements in their genome might be more susceptible to exogenous viral infections today, through, for example, interactions with immune pathways like the Herpes Simplex infection (KEGG:05168), eventually leading to excessive stimulation of brain immune responses such as increased presence of amyloid-beta protein.

However, our results should be interpreted with care. For example, the herpesvirus remnants mostly resembled the non-human envelope protein of the Chelonid α-herpesvirus. One can interpret this to have been infections of viruses similar to the Chelonid α-herpesvirus. However, envelope proteins are highly conserved within the virus kingdom, which might hint that the remnant stems from a different virus family. For example, a retrovirus, as they are much more present in the human genome compared to non-retroviral viruses. As another example, most virus signal was found within LTR repetitive elements according to the GRCh38 chromosomal locations. While this is not unexpected, given the mode of non-retroviral integration, it could also indicate retrotransposal or retroviral origin [8]. Nevertheless, all former research in the field of human paleovirology has been conducted on short-read sequencing data whereas our data can capture larger structural elements like virus integrations better, due to the nature of long-read sequencing data.

The findings regarding *Poxviridae* are new to the field of EVEs, and also need validation in other cohorts. Members of the *Poxviridae* family replicate inside the cytosol and thus chances of integrations are very low. Previously discovered EVEs, for example of the *Chuviridae*, were also thought not to integrate at all but were identified nevertheless [47]. It would be interesting to do similar experiments on vertebrate genomes other than human genomes to see if a poxvirus-like sequence can be identified as well.

Although our findings underscore the diversity and differences in the EVE landscape of individuals, we should note that we are highly biased to individuals with a Dutch background. Stronger support for our findings would be created if this study could be performed on data from various geographic populations.

In conclusion, we have shown that our genomes can be viewed as documents archiving the virus interactions we have had in the past. Our findings of an increased EVE presence in AD genomes with respect to CHC genomes ignites discussion on the infectious hypothesis, highlighting the importance of preventive measures against infectious diseases, demonstrating not only short-term benefits, but also long-term benefits for ourselves and possibly for our descendants.

## Data Availability

Sequencing data are available upon request from the corresponding authors.

## Consent

All participants and/or their legal representatives provided written informed consent for participation in clinical and genetic studies.

